# Identification of Risk Factors for Glaucoma Progression in Free-Text Clinical Notes using a Local Large Language Model

**DOI:** 10.1101/2025.09.26.25336746

**Authors:** Anshul Bhatnagar, Rafael Scherer, Gustavo A. Samico, Rohit Muralidhar, Naomi E. Gutkind, Vitoria Palazoni, Felipe A. Medeiros, Swarup S. Swaminathan

**Affiliations:** Bascom Palmer Eye Institute, University of Miami, Miami, FL, USA; Department of Ophthalmology and Visual Sciences, Escola Paulista de Medicina Universidade Federal de São Paulo, São Paulo, Brazil

**Keywords:** large language model, artificial intelligence, electronic health records, glaucoma, risk factors

## Abstract

**Purpose:** To evaluate the performance of a large language model (LLM) in identifying medication non-adherence, visit non-adherence, and family history of glaucoma (FHoG) in clinical notes from the electronic health record (EHR).

**Methods:** We extracted clinical notes of 1,250 glaucoma-related encounters between 2014 and 2024 and structured EHR family history field data from the Bascom Palmer Ophthalmic Repository, with 125 randomly selected notes (10%) used for prompt development and excluded from analysis. Two fellowship-trained glaucoma specialists labeled notes for evidence of non-adherence and FHoG. We utilized MedGemma-27B-text-it, a specialized medical LLM, to identify medication non-adherence, visit non-adherence, and FHoG. We calculated accuracy, sensitivity, and specificity of LLM performance for each task, Jaccard index for FHoG, and mean squared error (MSE) of number of family members with glaucoma.

**Results:** Prevalence of medication non-adherence, visit non-adherence, and FHoG were 7.3%, 4.7%, and 29.2%, respectively. LLM accuracy was 0.91 (sensitivity: 0.96; specificity: 0.91) for medication non-adherence and 0.96 (sensitivity: 0.97; specificity: 0.94) for visit non-adherence. For FHoG, LLM accuracy was 0.98 (sensitivity: 0.99; specificity: 0.99) with Jaccard index of 0.99, while EHR family history field accuracy and Jaccard index were 0.49 and 0.75, respectively. LLM and EHR MSE in quantifying the number of relatives with glaucoma were 0.05±0.56 and 0.85±1.80, respectively (p<0.001).

**Conclusions:** LLMs identified non-adherence to medication and visit schedules as well as degree of FHoG in clinical notes with high accuracy.

**Translational Relevance:** Local LLM pipelines can enable large-scale research into glaucoma risk factors that are unavailable in discrete EHR fields.

## Introduction

Glaucoma is a leading cause of irreversible blindness in the United States and abroad.^1,2^ Given glaucoma progresses asymptomatically, longitudinal monitoring and treatment are essential to prevent disease worsening. Key risk factors for progressive disease include older age, African ancestry, Latino ethnicity, elevated intraocular pressure (IOP), and thin central corneal thickness.^2^ Medication non-adherence, visit non-adherence, and family history of glaucoma (FHoG) are also significantly associated with glaucoma disease progression. Studies have shown that glaucoma patients who are lost to follow-up or unable to follow prescribed medication regimens have significantly poorer visual outcomes compared with treatment-adherent patients.^3–6^ Newman-Casey et al demonstrated that in a randomized clinical trial, glaucoma patients who were non-adherent to medications had a faster rate of visual field loss compared to adherent patients.^4^ Separately, genetic and familial studies have shown that a FHoG can drastically increase the risk of glaucoma and worse visual outcomes, and that this risk rises exponentially when more relatives are diagnosed with glaucoma. Patients with one sibling with glaucoma had an increased glaucoma incidence risk of 2.31, while those with four or more affected siblings with glaucoma had an increased incidence risk of 26.66.^7,8^

Electronic health record (EHR) systems have created a unique opportunity to develop large real-world datasets for research endeavors. EHR phenotyping refers to the concept of developing patient cohorts with a specific condition; additional extracted patient data can further characterize phenotypes. Data regarding specific glaucoma risk factors reside in discrete structured data elements in EHRs, such as age, self-reported ethnicity, tonometry, and pachymetry.^9–11^ These discrete data can be extracted efficiently, but key glaucoma risk factors such as medication and visit non-adherence are buried in unstructured clinical notes, complicating large-scale analysis. Information about these risk factors can currently only be gathered through manual chart review, which is error prone and impractical in larger clinical research initiatives.^12^ Although EHRs have formal medication lists or family history elements, research has repeatedly shown that such fields are often incorrect or outdated, and that unstructured clinical notes are often the most accurate source.^13–16^ Similarly, visit attendance data in EHRs is unreliable, as patient attendance is not well described by no-show percentages.

Prior studies have attempted to utilize natural language processing to study clinical notes, including work on extracting examination findings and diagnosis codes from clinician notes.^17^ Natural language models (NLM) are artificial intelligence algorithms that are trained not just to identify fields but also interpret text. Given that approximately 80% of all EHR data exists in an unstructured, narrative form,^18^ NLM models are becoming increasingly recognized for their ability to distill key information from free-text notes.^17^ Prior NLM research has focused on the use of large language models (LLMs), such as ChatGPT and BERT, which can effectively interpret ophthalmic information and free-text.^19^ Nonetheless, most LLMs face constraints in clinical settings involving protected health information (PHI), primarily because they require cloud-based processing and data sharing via public application programming interfaces. In countries with strict health privacy laws such as the United States, this limitation may preclude widespread adoption when analyzing clinical notes, as such text may contain PHI. Data security is paramount in such instances.

In contrast, a medium-sized LLM can run locally within a secure network, which is crucial when handling sensitive text.^20^ In this study, we aimed to develop a local LLM pipeline that could accurately determine whether glaucoma patients had a history of medication non-adherence, visit non-adherence, or FHoG per free-text clinical notes. Such a tool could not only help researchers rapidly extract these important risk factors from EHRs for real-world data research, but also facilitate risk-stratification of patients in a clinical setting.

## Methods

### Data Collection, Note Sampling, and Verification

This study was approved by the Institutional Review Board at the University of Miami. The requirement for informed consent was waived because of the retrospective nature of the study. The procedures and protocols followed during the study adhere to the Declaration of Helsinki and comply with the Health Insurance Portability and Accountability Act (HIPAA) for maintaining patient confidentiality and integrity.

This study used the Bascom Palmer Ophthalmic Repository (BPOR), which contains the data of over 70,000 patients evaluated for glaucoma at the Bascom Palmer Eye Institute (BPEI) in Miami, Florida.^21^ The database contains demographics, testing data, EHR-derived measurements, and clinical notes for included subjects. All data were extracted from Epic (Epic Systems, Verona, WI). We identified all signed clinical notes associated with an outpatient encounter that were written by a fellowship-trained glaucoma specialist between 2014 and 2024. Notes were only included if associated with an International Classification of Diseases (ICD)-10 glaucoma visit diagnosis (H40.X). Notes were excluded if the encounter type was hybrid, preoperative, postoperative, procedural, or consult given likely minimal detail regarding longitudinal glaucoma care. Notes that were fewer than 200 characters were also excluded. A total of 1,250 notes were randomly selected stratified by clinician to ensure equal representation of note styles. During note verification, eight notes were found to be attestations for trainee clinical visits and thus were excluded from further analysis. Only one note per subject was included. For each subject, the formal structured family history section was extracted directly from the EHR and filtered for mention of “glaucoma”. Notes were stored on a HIPAA-compliant secure Box server (Box, Redwood City, CA), which is authorized by the University of Miami to store PHI.

### Note Labeling and Prompt Engineering

A total of 125 randomly selected notes (10% of the full dataset) stratified by clinician were used to guide prompt engineering and were subsequently excluded from further analysis. These notes were used to create “few shot” examples, provide guidance regarding approaches to handling conflicting data, and generate a list of abbreviations observed in notes. Text of the three prompts is provided in the **Supplemental Material**.

Two fellowship-trained glaucoma specialists (GAS and NEG) labeled the remaining 90% of notes for evidence of current medication non-adherence (yes/no or not mentioned), visit non-adherence (yes/no or not mentioned), and FHoG (yes/no/not mentioned). For the non-adherence tasks, “not mentioned” was grouped with “no” (i.e., binary classification), as physicians would likely only document cases of non-adherence. For the FHoG task, “not mentioned” was considered as a distinct category under the assumption that providers would actively document both positive and negative FHoG. If a positive FHoG was documented, graders were asked to identify the family members with glaucoma, if discussed in the note. In cases of disagreement between the graders, a third fellowship-trained glaucoma specialist (SSS) served as an adjudicator. This process produced a ‘ground truth’ standard for each extraction task to subsequently assess model performance.

### Model Architecture, Setup, and Processing

We used MedGemma-27B-text-it, a specialized medical LLM developed by Google (Alphabet Inc., Mountain View, CA). This artificial intelligence model has been specifically trained to analyze medical text, such as clinician notes.^22^ To ensure no exposure of PHI, this LLM was installed and run on a local computer in a secure location at BPEI and housed within a virtual environment. All notes were processed with their corresponding encounter dates to provide temporal context. The model was implemented using the Hugging Face Transformers library with PyTorch backend. To maximize computational speed and efficiency, the model was configured to run with bfloat16 precision instead of float32. To ensure deterministic outputs, meaning that the model should return the same output when given the same input across devices or time, we disabled memory-efficient and flash self-attention mechanisms; we enabled mathematical self-attention instead.

We modified the key generation parameters (temperature=0.4, top_p=0.9, max_new_tokens=200) of the model to balance output consistency with appropriate variability. Generated outputs were parsed to extract structured JavaScript Object Notation (JSON) data with error handling for any malformed responses. The model was run without any formal training or fine-tuning to avoid significantly changing the native behavior of the model. Output tokens were reviewed, particularly in cases of false negatives and false positives, to provide insight into the model’s rationale for classification.

### Statistical Analysis

Model outputs were compared against the adjudicated standard for each note. To determine model performance for each data extraction task, accuracy, sensitivity, and specificity were calculated. For FHoG, overall accuracy was calculated across all three valid responses. The Jaccard index was calculated for FHoG to assess family member matching. The Jaccard index evaluates partial and exact matches between the predicted and true classifications, measuring similarity by dividing the size of their intersection by the size of their union. Regarding relatives with glaucoma, we converted the listed family members into a numeric estimate from the graded labels, LLM output, and EHR structured field. If the output was “Yes” with no family members listed, the value was listed as one. If the output included some indication of plurality (e.g., “multiple,” “cousins”) with no further details, the value was listed as two. We then calculated the mean squared error (MSE) of the LLM and EHR field when compared to the graded labels to determine their accuracy. Only subjects with a positive family history per the graded label were included in the MSE calculations to avoid overestimating accuracy of this task (i.e., an artificially low MSE estimate).

Gwet’s AC1 and the raw agreement rate were calculated to evaluate concordance rates between graders. Gwet’s AC1 is a useful assessment metric and alternative to Cohen’s kappa coefficient when prevalence of a condition is low. The Wilson score interval was used to calculate 95% confidence intervals (CI) for accuracy, sensitivity, and specificity. For Gwet’s AC1, 95% confidence intervals were estimated using bootstrap resampling with 1,000 iterations. MSE of LLM versus EHR structured field assessments were compared using the nonparametric Wilcoxon signed-rank test. A p-value <0.05 was considered statistically significant. All statistical analyses were conducted using Stata version 18 (StataCorp, College Station, TX, USA).

## Results

A total of 1,117 glaucoma-related encounter notes were used to analyze model performance. The LLM was able to successfully evaluate all notes without computational error. Prevalence of medication and visit non-adherence was 7.3% (n=81) and 4.7% (n=53), respectively, based on graded labels. Clinicians documented FHoG in 29.2% (n=326) of all subjects. Inter-rater reliability was excellent across all assessments. For medication non-adherence, Gwet’s AC1 was 0.914 (95% CI, 0.887–0.936) with a raw agreement of 95.7%. For visit non-adherence, the AC1 was 0.961 (95% CI, 0.943–0.977) with a raw agreement of 98.0%. For FHoG, the AC1 was 0.913 (95% CI, 0.886–0.928) with a raw agreement of 93.5%.

For classifying medication non-adherence, the LLM had an accuracy of 0.91 (95% CI: 0.89-0.93), with sensitivity and specificity of 0.96 (95% CI: 0.90-0.99) and 0.91 (95% CI: 0.89-0.92), respectively (**Figure 1**). For classifying visit non-adherence, the LLM had an accuracy of 0.96 (95% CI: 0.95-0.97), with sensitivity and specificity of 0.97 (95% CI: 0.95-0.97) and 0.94 (95% CI: 0.84-0.98), respectively (**Figure 2**).

**Figure 1.**
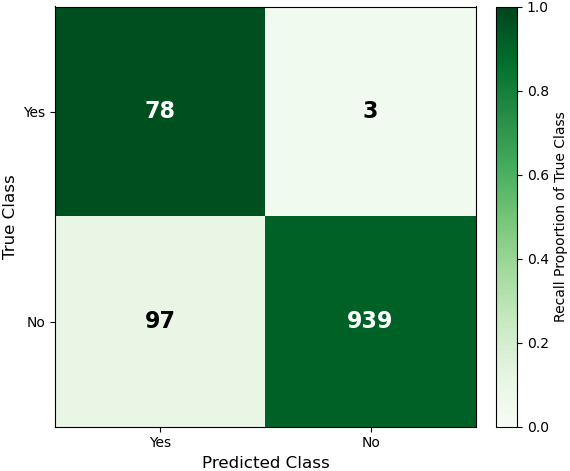
Confusion matrix for medication non-adherence. Classification was binary (“yes” or “no / not mentioned”).

**Figure 2.**
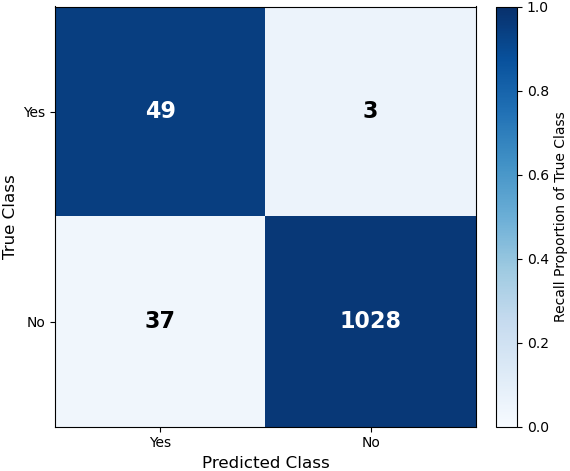
Confusion matrix for visit non-adherence. Classification was binary (“yes” or “no / not mentioned”).

For FHoG, LLM accuracy was 0.98 (95% CI: 0.97-0.99; **Figure 3A**), with sensitivity of 0.99 (95% CI: 0.98-0.99) and specificity of 0.99 (95% CI: 0.98-0.99). In terms of family member matching, the Jaccard index was 0.99 (95% CI: 0.98-0.99). In contrast, the structured field for family history in the EHR had an accuracy of only 0.49 (95% CI: 0.46-0.52; **Figure 3B**), significantly lower than that of the LLM (p<0.001). Sensitivity and specificity of the EHR structured field were 0.76 (95% CI: 0.71-0.80) and 0.81 (95% CI: 0.78-0.83), respectively. The Jaccard index was 0.75 (95% CI: 0.73-0.78).

**Figure 3.**
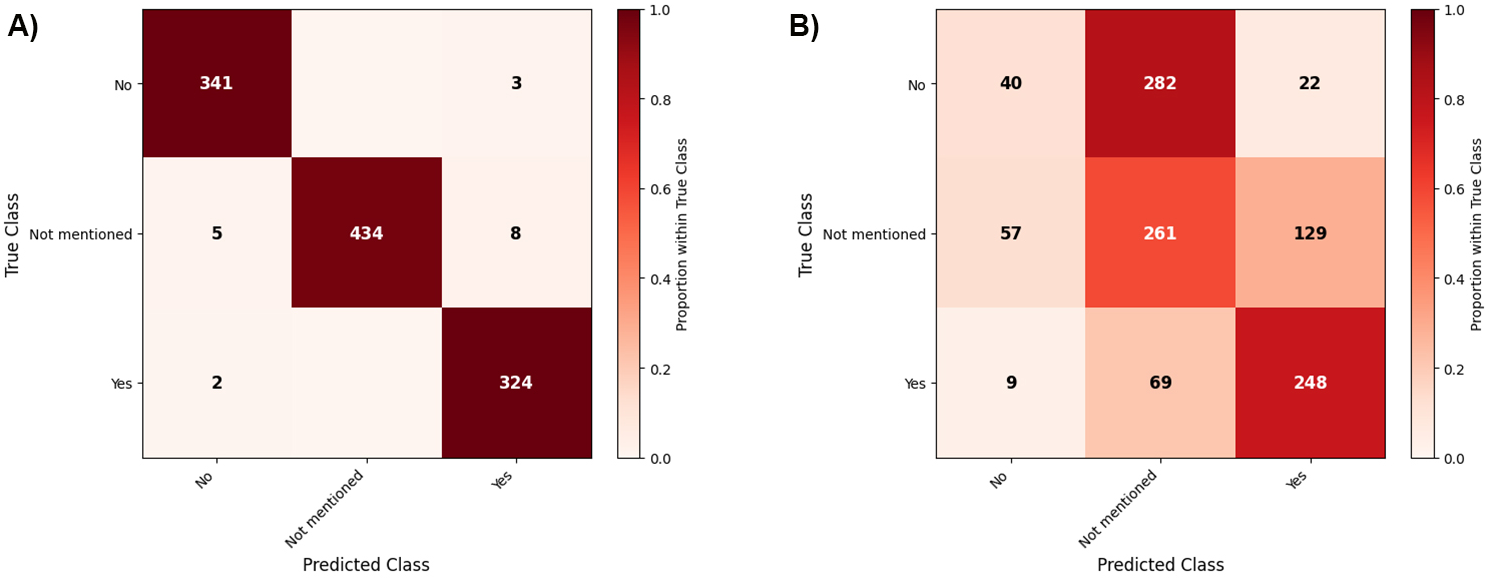
Confusion matrix for family history of glaucoma using A) large language model classification and B) electronic health record structured field classification. Classification was ternary (“yes”, “no”, or “not mentioned”).

According to the clinicians’ assessment, subjects with positive FHoG had a median of one affected relative (range: 1-4; **Figure 4**). The MSE of the LLM when estimating the number of relatives with glaucoma was 0.05±0.56. The MSE of the EHR structured field was much larger, 0.85±1.80 (p<0.001). In 2.1% of notes, the graded label noted a negative FHoG but the subject was noted to have a positive FHoG per the structured EHR field. In 11.5% of notes, the graded label for FHoG in the clinical note was “not mentioned” but the subject had a positive FHoG per the structured EHR field. **Table 1** contains all performance metrics of all models for comparison.

**Figure 4.**
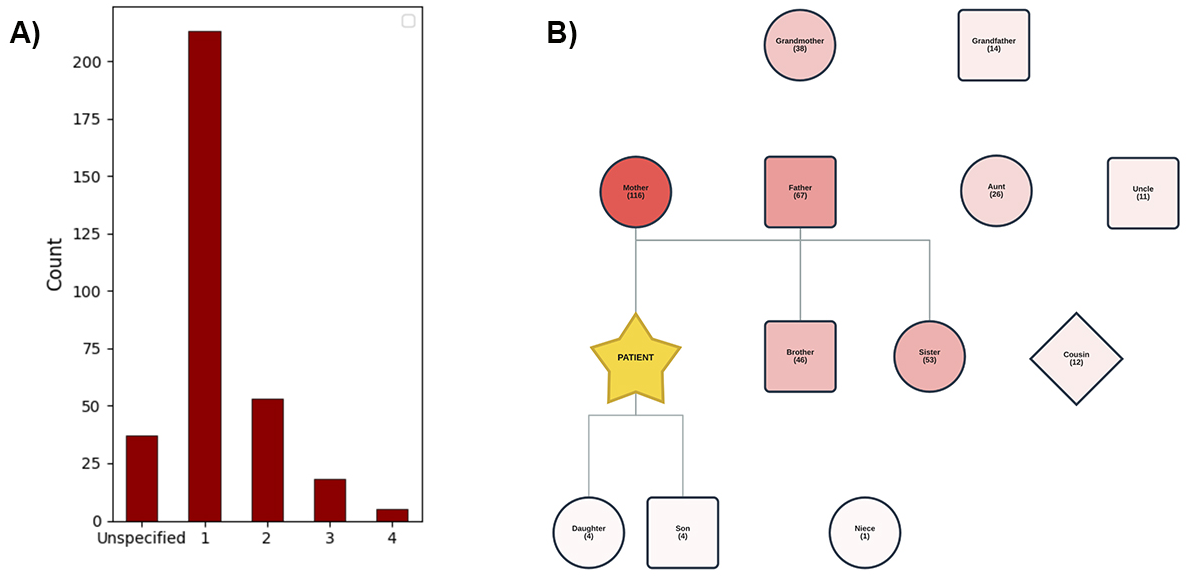
A) Histogram describing the distribution of the number of family members with glaucoma per graded labels. B) Diagram reflecting the frequency of relatives noted to have glaucoma per the graded labels. Darker colors reflect higher frequency (count indicated in parentheses).

**Table 1.** Summary of large language model performance metrics for classification tasks with 95% confidence intervals.

| **Metric** | **Medication**<br>**Non-**<br>**Adherence** | **Visit**<br>**Non-Adherence** | **FHoG** | **FHoG (EHR**<br>**Structured Field)** |
| --- | --- | --- | --- | --- |
| *Accuracy* | 0.91 (95% CI: 0.89–0.93) | 0.96 (95% CI: 0.95–0.97) | 0.98 (95% CI: 0.97–0.99) | 0.49 (95% CI: 0.46–0.52) |
| *Sensitivity* | 0.96 (95% CI: 0.90–0.99) | 0.97 (95% CI: 0.95–0.97) | 0.99 (95% CI: 0.98–0.99) | 0.76 (95% CI: 0.71–0.80) |
| *Specificity* | 0.91 (95% CI: 0.89–0.92) | 0.94 (95% CI: 0.84–0.98) | 0.99 (95% CI: 0.98–0.99) | 0.81 (95% CI: 0.78–0.83) |
| *MSE* | - | - | 0.05 ± 0.56 | 0.85 ± 1.80 |

EHR: Electronic Health Record, FHoG: Family History of Glaucoma, MSE: Mean Squared Error

## Discussion

We demonstrate in this study that a tailored, secure, locally-installed LLM pipeline can perform key extraction tasks from EHR-derived clinical notes with high accuracy. The model was 91% accurate at identifying medication non-adherence, 96% accurate at identifying visit non-adherence, and 97% accurate at detecting FHoG. These risk factors are associated with worse glaucoma outcomes but have traditionally been difficult to study in glaucoma research due to their inaccessibility. By converting unstructured, narrative information to discrete, structured data that can be batch-extracted and analyzed, LLMs have the potential to aid not only research endeavors but also identify high-risk glaucoma patients in large practice settings. To our knowledge, this work represents the first study to develop a locally-installed LLM pipeline that identifies and interprets risk factor data from free-text clinical notes in ophthalmology.

The LLM was highly sensitive and specific (0.96 and 0.91, respectively) for the detection of medication non-adherence within clinical notes. Similarly, the model performed with high sensitivity and specificity (0.97 and 0.94, respectively) in identifying visit non-adherence in clinical notes. **Table 2** contains examples of false positive and false negative misclassifications for both medication and visit non-adherence. Medication adherence misclassification appeared to be due to etiologies such as difficulty in interpreting past versus current non-adherence, lack of clarity regarding whether a patient self-discontinued a medication due to side effect or without reason, or an elevated IOP with a discussion of the importance of compliance in the clinician’s note. Visit adherence misclassification was often due to misinterpretations regarding changes in clinical providers, missed follow-up with non-glaucoma clinicians, or lack of clarity regarding where the patient was supposed to follow-up (e.g., at BPEI or with an external provider). In contrast to FHoG classification, the non-adherence tasks required greater interpretation of note text, naturally leading to reduced accuracy (**Table 1**). While false positives were more common than false negatives, a false positive (i.e., declaring non-adherence when patient was adherent) is likely less consequential than a false negative (i.e., declaring adherence when patient was non-adherent). To this end, the sensitivities of these models remain robust, reflecting low false negative rates. One could also argue that the false positives in each task may actually identify patients whose actions suggest potential adherence challenges (e.g., forgetting use of drops on day of clinic visit), albeit they were not formally classified as non-adherent by graders.

**Table 2.**
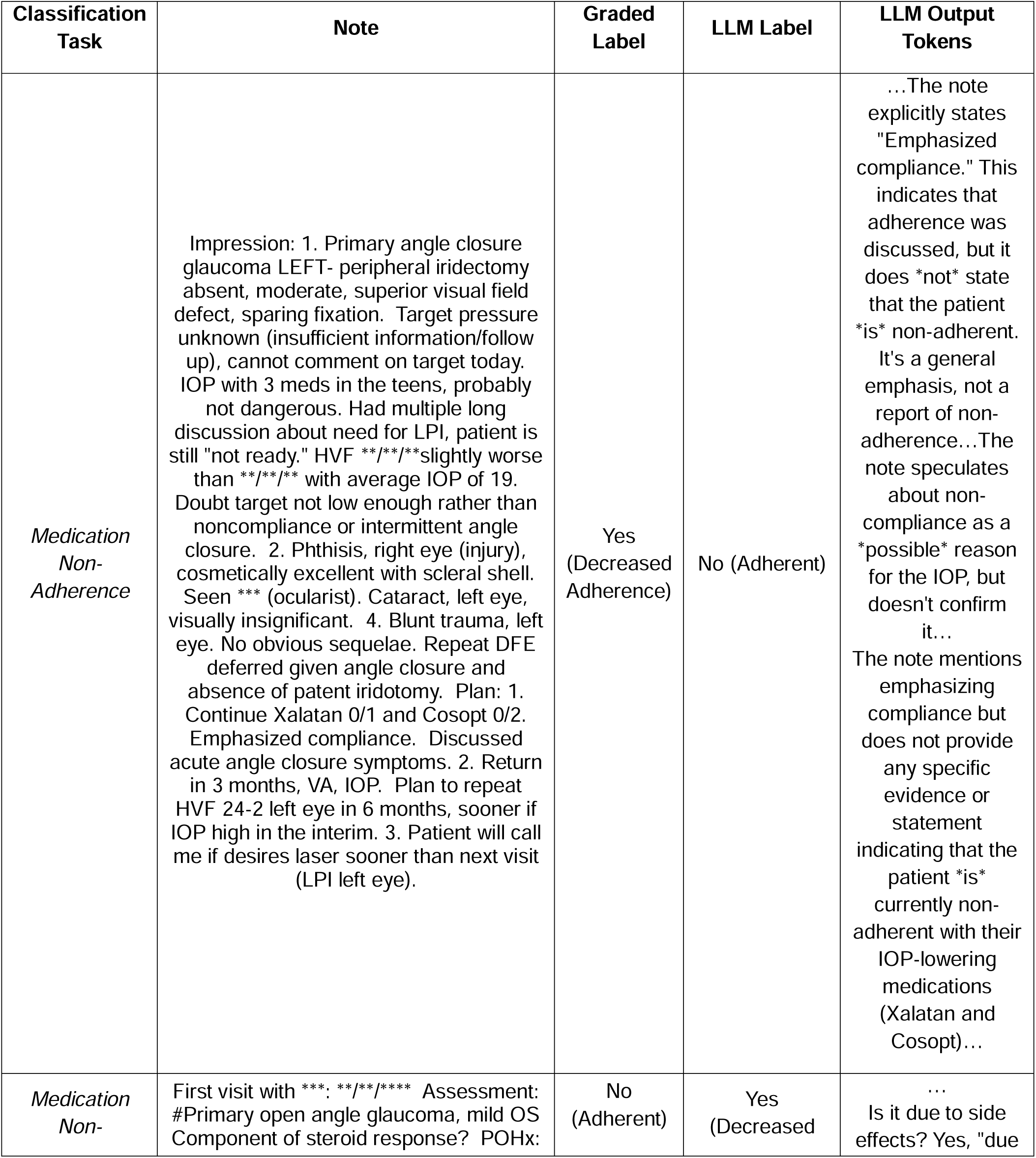
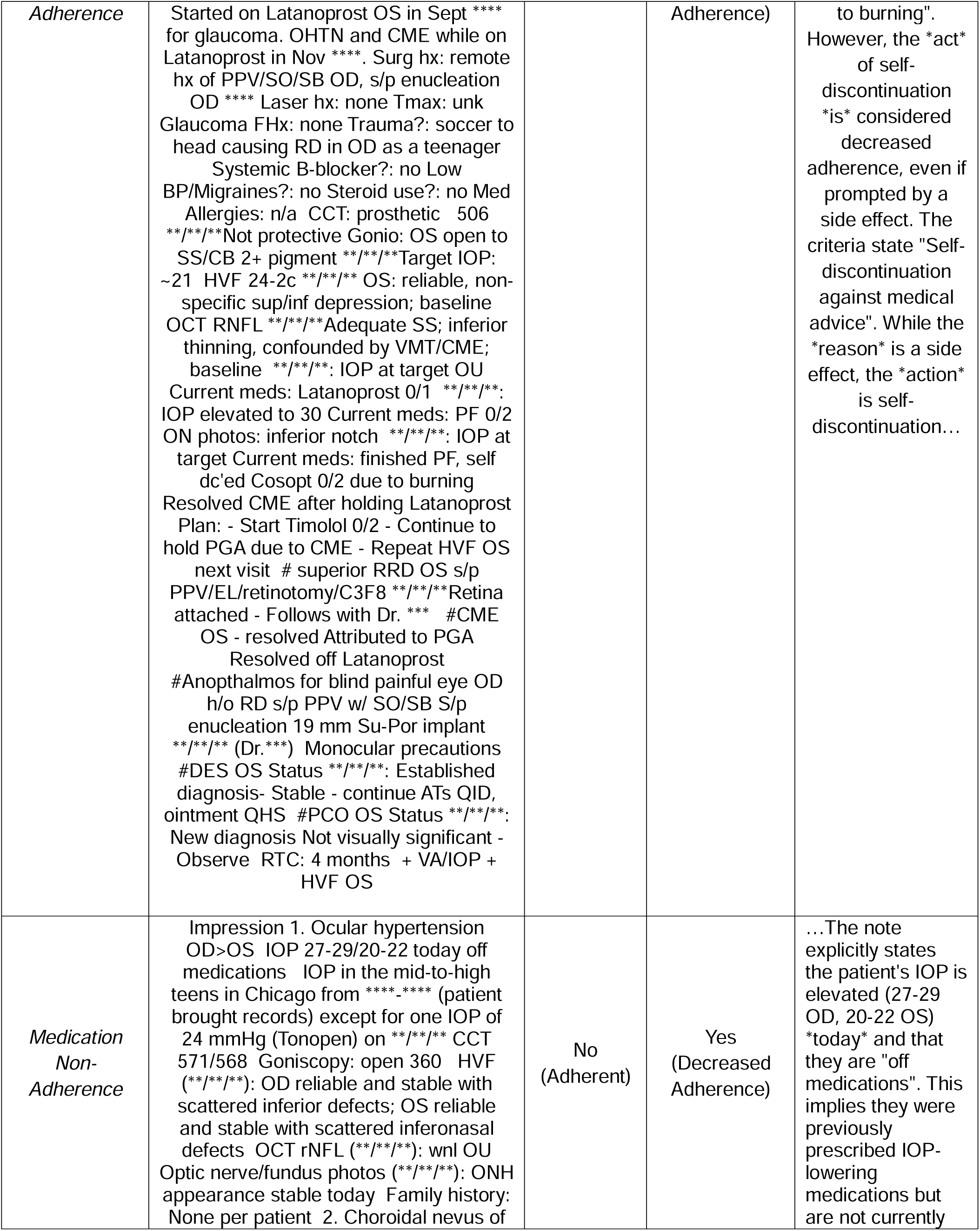
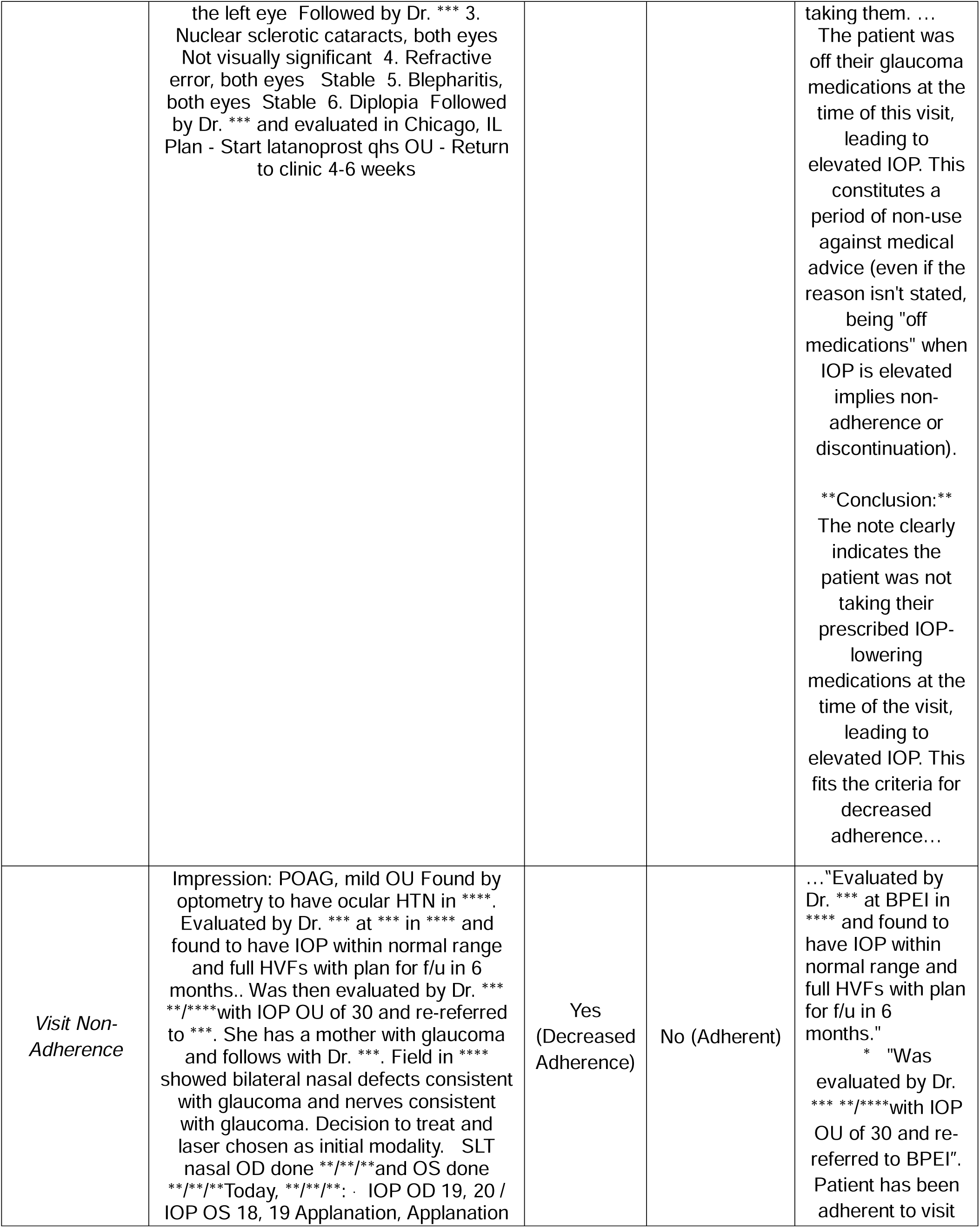
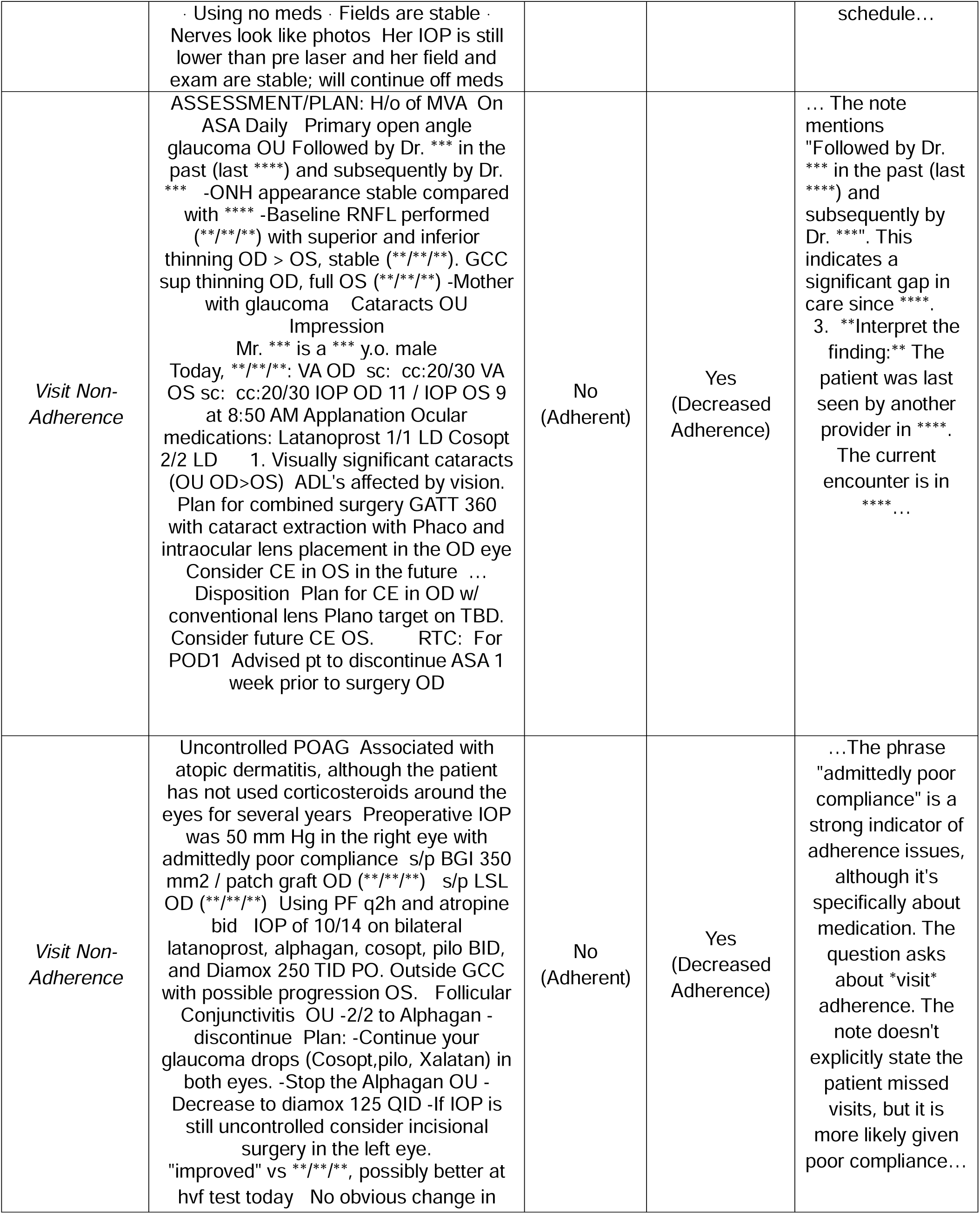
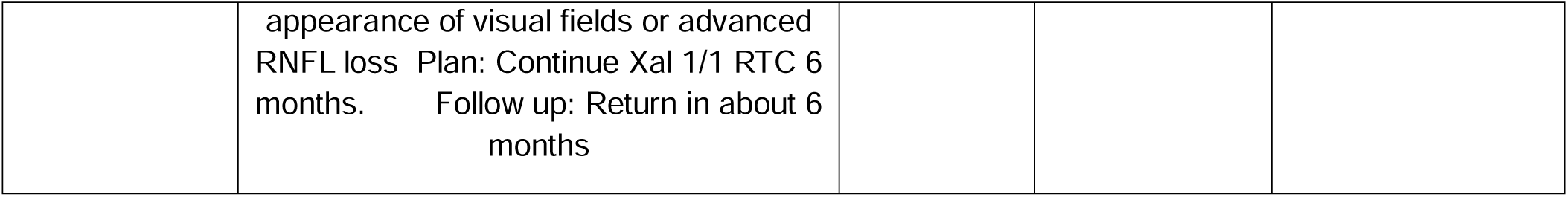
Examples of medication and visit non-adherence misclassification by the large learning model (LLM). The output tokens provide insight into the rationale for decision-making by the LLM. Any names, dates, or other potentially identifying information have been replaced by asterisks.

Medication non-adherence is challenging to assess using EHR medication lists, as these fields are often inaccurate for ophthalmic medications. One study of glaucoma patients demonstrated that a third of all patients had at least one glaucoma medication mismatch between the formal EHR medication list and the clinical text note.^15^ These findings suggest that ophthalmologists and optometrists do not update EHR medication lists frequently, despite 90% of ophthalmologists acknowledging the importance of assessing glaucoma medication adherence during clinic visits.^23^ Similarly, claims databases may not accurately reflect medication adherence, as patients may fill prescriptions as requested but still use medications incorrectly.^24^ Consequently, free-text clinical notes are likely the most accurate source for such information.

Visit non-adherence is another significant risk factor for glaucoma progression that is often understudied and challenging to track through structural mechanisms in EHRs. Although raw no-show percentage (number of no-show visits / number of scheduled clinic visits), can be extracted from EHRs, this statistic does not always accurately reflect true visit adherence. Such percentages can be deceivingly low. For example, glaucoma patients that are lost to follow-up for years after missing just one visit will have a no-show percentage that does not reflect their extended lack of follow-up, which puts them at significant risk of disease progression.^25^ Second, such statistics are often calculated across multiple specialties in a hospital system within the EHR and thus may not be specific to glaucoma visit adherence, which can be significantly decreased compared to that of other ophthalmic subspecialties.^26^ Again, clinic notes may serve as the most accurate source of information regarding visit adherence specific to glaucoma care. Analysis of these additional patient dimensions via LLMs could facilitate further characterization of glaucoma phenotypes in EHR data.^27^ In addition, larger organizations could create registries of high-risk glaucoma patients identified using such an LLM pipeline. These individuals may benefit from specialized and targeted interventions to improve visit adherence, such as personalized letters, telephone reminders, and social worker support.^28,29^ Use of these registries could lead to improved visual outcomes for patients that might otherwise be lost to follow-up in larger practices.

FHoG is another key risk factor for glaucoma progression that is purportedly tracked within structured family history EHR fields. While such fields can be easily extracted, prior work in neurology and obstetrics/gynecology has shown that such lists are often inaccurate and infrequently updated.^14,30^ Our study confirmed that with respect to glaucoma, EHR structured family history data were accurate less than half the time. In contrast, only 13.6% of patients had a positive FHoG per the structured EHR field that was missed in the clinical note (labeled as “no” or “not mentioned”). It is important to note that since the clinical note was treated as the ground truth, the LLM output had a potential advantage over the EHR field, which could make such comparisons challenging. However, given ophthalmologists and optometrists typically use their notes to track relevant family history details and are less likely to use the structured EHR field, our findings suggest that the EHR field may not be the optimal source of eye-related family history, particularly for research purposes.

In contrast, the LLM model exhibited high sensitivity and specificity (0.99 for both) in detecting FHoG from clinical notes. While the non-adherence tasks required more interpretation, FHoG assessment was likely an easier task involving the identification and reporting of relevant information, leading to strong model performance (**Table 1**). When assessing the number of family members with glaucoma, the LLM maintained excellent accuracy (MSE 0.05±0.56). This finding is particularly important given that the risk of glaucoma rises exponentially with the number of affected relatives with glaucoma.^8^ LLMs may allow for accurate, quantified assessments of FHoG, which can provide valuable details for research work in glaucoma genetics or disease progression assessments. EHR glaucoma phenotypes could potentially incorporate quantification of FHoG to yield more specific cohorts.

NLMs have become increasingly popular in medicine for their ability to interpret free narrative text, although their use thus far for ophthalmic purposes has been limited.^17^ Prior ophthalmic research has used NLMs to predict glaucoma progression, identify cataract surgery complications, determine slit lamp-examination findings, and recognize ophthalmic diagnoses from clinic notes.^17,31–36^ However, fewer studies have used NLMs to examine risk factors for glaucoma progression. Lin et al. used a natural language processing tool to evaluate if medication adherence was mentioned in clinical notes.^15^ However, this smaller study differs from our analysis as it solely identified the presence of adherence information; no further interpretation of the adherence text was pursued.

In contrast to the focus on cloud-based LLMs in medical research, our use of a nimble, locally installed LLM provides various advantages. Most critically, the latter can be operated on a local network without the need for cloud architecture.^20^ LLMs often require data to be shared across networks via third-party application programming interfaces. Many countries such as the United States have strict laws regarding PHI, which could limit widespread use of cloud-based LLMs in clinical or health research processes.^37^ Our study demonstrated that a locally-installed LLM can achieve excellent performance in such tasks, even without extensive model training. Studies traditionally use a train/test approach to evaluate model performance, but accurate assessments can be challenging when data are sparse and prevalence of the target condition is low.^38–40^ In this work, we utilized a “few-shot” approach, with 10% of notes used for prompt engineering and the remainder utilized to evaluate model performance. A local LLM pipeline is uniquely suited to implementation within clinical workflows when working with sensitive data.

Our study has natural limitations. First, the notes analyzed in this study were from one institution, which may limit the generalizability of our LLM pipeline. However, it is important to be mindful of institution-specific writing styles and abbreviations in clinical notes, which may limit the accuracy of using the same prompt at different institutions. Rather, our work suggests that individual institutions may need to follow a similar “few-shot” approach to capture institution-specific writing patterns. This work and the prompts provided in this study may serve as a foundation for initial testing at different institutions with further prompt engineering. Future work could also involve extracting a standardized output from a LLM pipeline run locally at individual institutions, which could be incorporated into larger multi-institutional studies regarding glaucoma progression. Second, the ideal LLM can only be as accurate as clinician notes, which may miss key information discussed during patient encounters.^41,42^ If clinicians are prone to under-reporting adherence challenges in their notes, the LLM will follow suit.

This study demonstrated that a domain-adapted, locally-installed LLM pipeline can identify medication non-adherence, visit non-adherence, and FHoG including quantification of family members with high accuracy. Data regarding these risk factors for glaucoma progression do not reside in structured EHR fields that can be easily extracted, making them challenging to include in glaucoma data science research. Implementation of a local, secure LLM, which offers many healthcare-specific advantages, may allow for rapid extraction and interpretation of unstructured data, potentially even supporting the creation of novel variables in large public datasets. Local LLM pipelines have the potential to improve how clinical researchers utilize sensitive free text to extract valuable data and to potentially support targeted patient interventions in clinical settings.

## Data Availability

All data produced in the present study are available upon reasonable request to the authors.

## Acknowledgements

None

## Meeting Presentation

Submitted for presentation at American Glaucoma Society Annual Meeting 2026, Association for Research in Vision and Ophthalmology Annual Meeting 2026

